# Pathophysiological mechanisms of uterine niches (cesarean scar defects) associated with abnormal bleeding and infertility: a systematic review

**DOI:** 10.1101/2025.07.10.25331318

**Authors:** Gabriel M. Lino, Pauliana V. M. Galvão, Maria L. F. da Silva, Priscila Maria de Barros Rodrigues, Valda L. M. Luna, Marcos Cezar Feitosa de Paula Machado, George A. M. Conrado

## Abstract

**Aim:** To review the pathological findings of uterine niches in women presenting with infertility and abnormal uterine bleeding.

**Methods:** The Medline, Embase, and Cochrane Central databases were searched until 1 May 2025. We included cohorts, case-controls, and case series. The risk of bias was evaluated with ROBINS-I, and the certainty of evidence was presented according to GRADE.

**Results:** We included fourteen studies (637 women). Chronic inflammation was observed in 54% (95% CI 25 to 81%; 5 studies, 280 women; I^2^ = 94.3%); fibrosis in 83% (95% CI 51 to 96%; 6 studies, 266 women; I^2^ = 92.1%); endometrial defects in 90% (95% CI 75 to 96%; 4 studies, 177 women; I^2^ = 64.6%); ectopic endometrium in 26% (95% CI 18 to 36%; 9 studies, 411 women; I^2^ = 74.2%); and vascular remodeling in 81% (95% CI 75 to 86%; 6 studies, 205 women; I^2^ = 0.0%). There was an increased risk of endometrial defects in women with niches compared to those with prior cesarean deliveries (RR 2.13, 95% CI 1.27 to 3.55; 2 studies, 124 women; I^2^ = 0.0%), similarly for ectopic endometrium (RR 2.86, 95% CI 1.03 to 7.96; 2 studies, 97 women; I^2^ = 0.0%). Additionally, niches exhibited a pro-inflammatory phenotype, characterized by elevated expression of CD138 and TNF-α.

**Conclusions:** The observed pathological features suggest an underlying process driven by the interplay of chronic inflammation, altered immune cell recruitment, fibrosis, and vascular remodeling within the regenerating endometrium.

**Registration number:** CRD420251049667.

## INTRODUCTION

Long-term morbidity following cesarean delivery has become a major concern in obstetrics ^1^. Reduced fecundability, increased risks of placenta accreta, placenta previa, and abnormal uterine bleeding have all been associated with prior cesarean ^1,2^. However, the mechanisms underlying these associations are extremely unclear. Over the past decade, growing attention has been directed toward anatomic defects at the site of the uterine scar, commonly referred to as uterine niches or cesarean scar defects, as key contributors to these adverse outcomes.

Uterine niches are present in 50 to 60% of women who have had a cesarean delivery ^3^, classically associated with abnormal uterine bleeding and infertility, although dysmenorrhea and dyspareunia are also often reported ^4^. Crucially, these symptoms can persist for years after the surgery.

In theory, damage to the endometrium and endometrial-myometrial interface is one of the promoters of niche pathogenesis ^5^. Thereafter, local dysfunctional hormonal, metabolic, and inflammatory mechanisms lead to reproductive disorders ^6^. It is assumed that the presence of extensive fibrosis and ectopic endometrium promotes symptom onset. However, the exact mechanisms are yet unknown. Such an understanding could provide valuable insights into both clinical management and prevention, aimed at reducing long-term adverse reproductive outcomes following cesarean delivery. Therefore, this review aimed to systematically review pathological findings in symptomatic uterine niches.

## METHODS

This systematic review was registered prospectively in the PROSPERO database (CRD420251049667). We followed the PRISMA 2020 guidelines for the reporting of systematic reviews and meta-analyses ^7^.

### Criteria for considering studies for this review

We considered randomized controlled trials, cohorts, case controls, and case series reporting pathological findings of women diagnosed with uterine niches after cesarean delivery and presenting with abnormal uterine bleeding, infertility, or related disorders. The following outcomes were sought in these studies: inflammation (and inflammatory markers), fibrosis, endometrial defects, presence of ectopic endometrium, asynchrony of the menstrual cycle in the scar, and angiogenesis or changes in the surrounding vasculatures (including dilated capillaries and thick-walled vessels).

### Search methods for identification of studies

We searched the Medical Literature Analysis and Retrievel System Online (Medline), Embase, and the Cochrane Central Register of Controlled Trials (Central) databases until 1 May 2025. A combination of the following terms, and variations, composed the search query: “cesarean scar defect”, “cesarean scar”, “uterine scar”, “uterine defect”, “isthmocele”, “uterine niche”, and “histology”. The search was adapted for each database. There were no language or publication date restrictions.

### Data collection and analysis

Two investigators were responsible for independently screening the title and abstract of the identified records. In this stage, if one of the authors considered the record eligible, both assessed the full text. Subsequently, they proceeded with the selection and extraction of key information from the included studies into a previously cross-validated file. Disagreements on which studies should be included in the review were resolved by discussing with a third investigator. Extracted information included authorship, conflict of interest, funding, country, setting, population, inclusion and exclusion criteria, information on baseline characteristics, adjustment methods for baseline imbalances, outcomes reported and how they were assessed, follow-up of participants, information on risk of bias, suture techniques in uterine closure, histological observations, and immunohistochemistry markers of inflammation, proliferation, and hormone expression (CD138, CD16, CD3, CD20, CD56, CD68, CD163, myeloperoxidase [MPO], tryptase, tumor necrosis factor-α [TNF-α], α-smooth muscle actin [α-SMA], Ki67, CD10, CD31, estrogen receptor [ER], and progesterone receptor [PR]).

Specific definitions for the outcomes were not required for inclusion in the review, as long as they were reported in the histological analyses. Uterine niches were considered appropriately defined when described as defects in the cesarean scar measuring more than 2 mm. When clear definitions for uterine niche diagnosis or outcome classification were lacking, the available information was recorded and used to inform the risk of bias assessment.

### Risk of bias assessment in included studies

Two investigators independently used the Cochrane risk-of-bias tool for nonrandomized studies of intervention (ROBINS), version 2024, to assess the risk of bias/quality of the included studies ^8^. When different classifications were reached, the final risk assessment was reached by discussion.

### Synthesis methods

We used an inverse-variance random effects model, reporting dichotomous data as Relative Risk (RR) with 95% Confidence intervals (CI) and continuous outcomes as mean differences (MD) and 95% CI. We also presented the baseline risk and risk difference (RD) for the critical outcomes ^9^. Statistical heterogeneity was assessed by calculating tau, tau^2^, and I^2^. No specific threshold of I^2^ was considered a significant indicator of heterogeneity; rather, a contextualized I^2^ was considered based on visual inspection and acceptable between-study estimate variation ^10^. We investigated publication bias with funnel plots.

The analyses were conducted in R, with the package metafor ^11^.

### Certainty of the evidence assessment

The overall certainty of the evidence for each outcome was assessed by two investigators using the GRADE approach, formulating a summary of findings and certainty of evidence tables containing the critical outcomes investigated. There were no disagreements; this process was conducted by deliberation.

The Grading of Recommendations, Assessment, Development and Evaluation (GRADE) handbook was followed to determine the quality of the evidence of the analyses conducted ^12^. For the evaluation of imprecision, the update provided in 2022 was followed ^13^. Inconsistency was evaluated by investigating contextualized heterogeneity, based on whether reasonable explanations for study variations could be provided ^10^.

The tables were generated using the GRADEpro GDT software available online.

## RESULTS

### Results of the search

We identified 447 records in three databases, Medline, Embase, and Central (Figure 1). After excluding duplicates and records marked as ineligible (case reports and reviews), we assessed the title and abstract of 318 records. Subsequently, we excluded 292 records for being outside the review scope. One additional study was excluded because the digital record was not available. The full text was extracted from 25 records. Finally, we excluded eight studies that did not report any histological analysis and three that did not investigate uterine niches.

**Figure 1.**
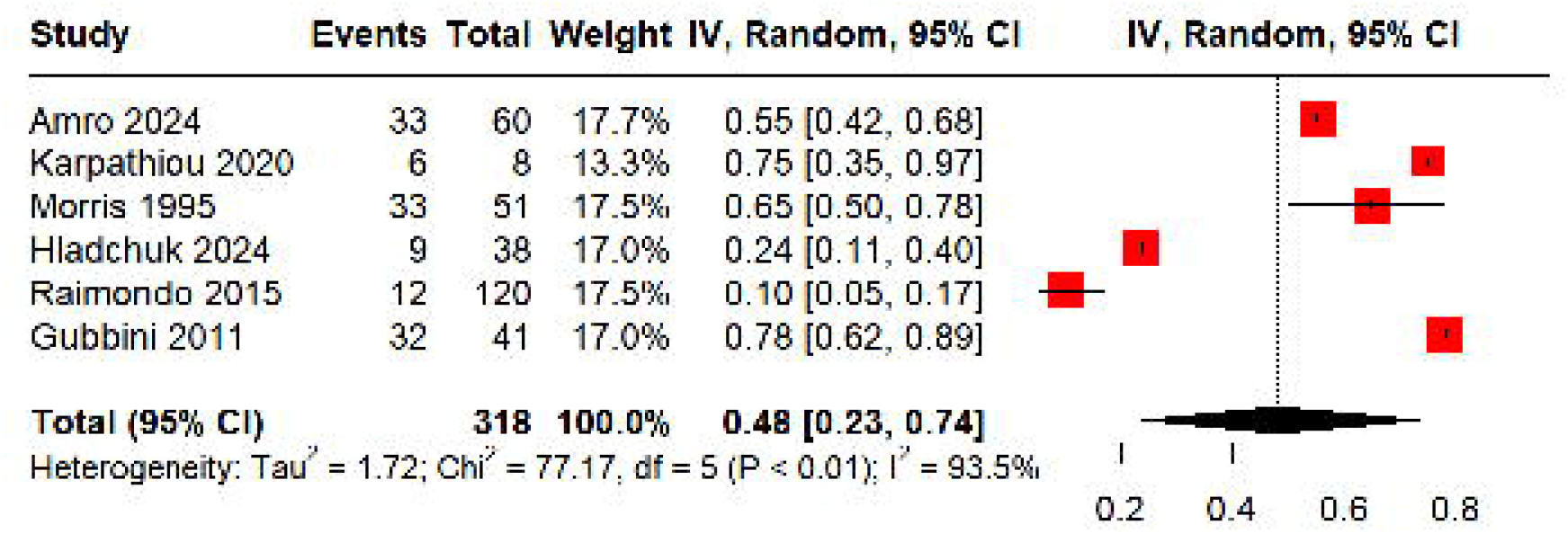
Legend PRISMA (Preferred Reporting Items for Systematic Reviews and Meta-Analyses) 2020 flow diagram of study selection.

### Included studies

Fourteen studies met the inclusion criteria for this review. Their main characteristics are summarized in Table 1, with additional details provided in Table S1. Additionally, the most common symptoms reported were abnormal uterine bleeding and infertility. However, in some cases, these symptoms could not be clearly attributed to the uterine niche due to coexisting comorbidities – an issue further addressed in the risk of bias assessment.

Histological samples were obtained either through hysteroscopic resection or hysterectomy, with varying time intervals between symptom onset and surgical management.

Commonly reported histological features included chronic inflammation, fibrosis, endometrial defects, ectopic endometrium, asynchronous menstrual cycle, vascular remodeling, and immunohistochemical markers of inflammation and cell proliferation. A visual summary of these findings across studies is provided in Figure S1.

Most studies employed single-arm, retrospective, or prospective designs, typically involving nonconsecutive patient series.

### Risk of bias

The overall risk of bias is presented in Table 1, while a more detailed description of each domain is provided in Table S2. In general, the quality of most studies was limited by selection bias and unclear reporting of pathological findings (reporting bias). Except for Wang 2025, Raimondo 2015, and Gubbini 2011, inadequate adjustment for baseline imbalances and nonconsecutive patient recruitment led to substantial residual confounding, resulting in at least a serious risk of bias for all other studies.

### Analyses of histopathological observations

#### Chronic inflammation

Six studies investigated chronic inflammation, resulting in a pooled prevalence of 48% (95% CI 23 to 74%; 6 studies, 318 women; I^2^ = 93.5%; very low-certainty evidence; Figure 2). Excluding the only study at critical risk of bias, the prevalence changes to 54% (95% CI 25 to 81%; 5 studies, 280 women; I^2^ = 94.3%; very low-certainty evidence; Figure S2). Raimondo 2015 had a conflicting estimate with the remaining studies; the pooled prevalence further changed to 66% when it was excluded (95% CI 54 to 76%; 4 studies, 160 women; I^2^ = 48.9%; very low-certainty evidence).

**Figure 2.**
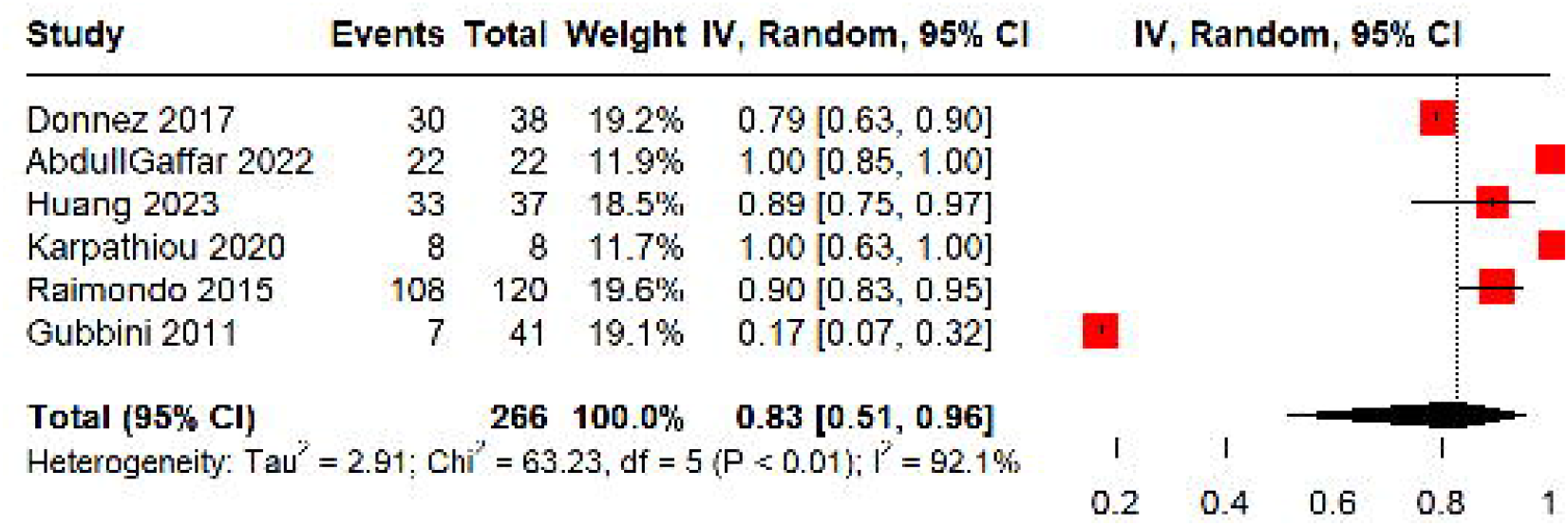
Legend Proportion of chronic inflammation in histopathological observations in women with symptomatic uterine niches. IV, inverse variance.

#### Fibrosis

Six studies investigated fibrosis, with a pooled prevalence of 83% (95% CI 51 to 96%; 6 studies, 266 women; I^2^ = 92.1%; very low-certainty evidence; Figures 3 and S3). As in the previous outcome, Gubbini 2011 and Raimondo 2015 had conflicting results. Excluding Gubbini from the fibrosis analysis results in a prevalence of 88% (95% CI 81 to 93%; 5 studies, 225 women; I^2^ = 24,3%; low-certainty evidence).

**Figure 3.**
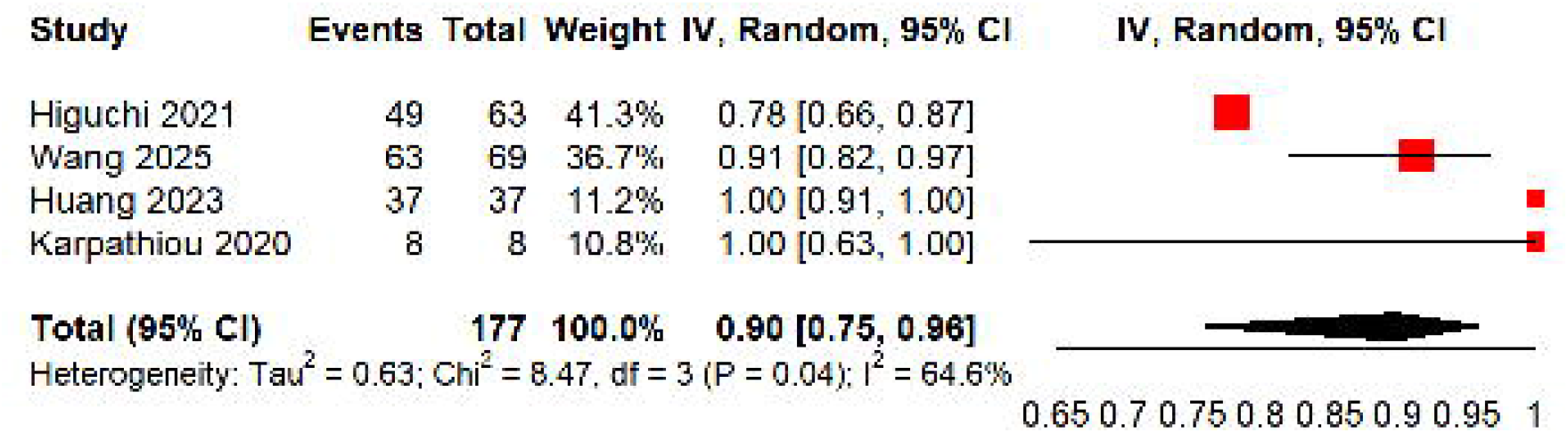
Legend Proportion of fibrosis in histopathological observations in women with symptomatic uterine niches. IV, inverse variance.

#### Endometrial defects

Four studies reported endometrial defects, resulting in a prevalence of 90% (95% CI 75 to 96%; 4 studies, 177 women; I^2^ = 64.6%; very low-certainty evidence; Figure 4). Additionally, Huang 2023 and Higuchi 2021 observed an increased risk of endometrial defects in women with niches compared to those with prior cesarean deliveries but without niches (RR 2.13, 95% CI 1.27 to 3.55; 2 studies, 124 women; I^2^ = 0.0%; low-certainty evidence; Figure S4). Further, Wang 2025 also observed an increased risk of endometrial defects in women with symptomatic niches compared to asymptomatic niches (RR 2.71, 95% CI 2.00 to 3.67; 1 study, 155 women; low-certainty evidence).

**Figure 4.**
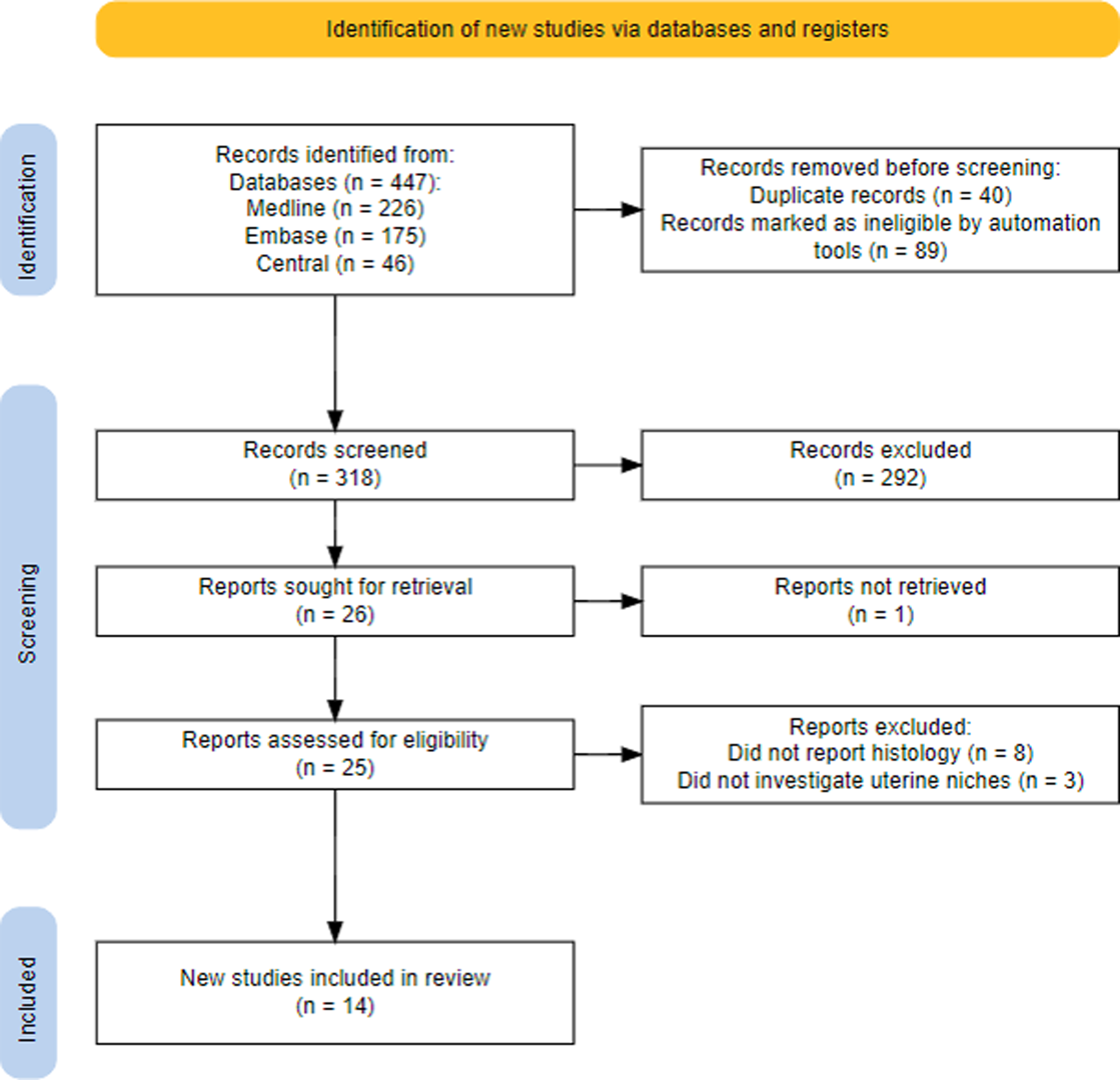
Legend Proportion of endometrial defects in histopathological observations in women with symptomatic uterine niches. IV, inverse variance.

#### Ectopic endometrium

Ten studies reported the presence of ectopic endometrium within the scar, with a pooled prevalence of 31% (95% CI 20 to 44%; 10 studies, 449 women; I^2^ = 84.8%; very low-certainty evidence; Figure S5). Excluding studies at critical risk of bias resulted in a prevalence of 26% (95% CI 18 to 36%; 9 studies, 411 women; I^2^ = 74.2%; very low-certainty evidence). Additionally, two studies compared women with uterine niches to those with prior cesarean deliveries but without niches. The presence of ectopic endometrium was associated with an increased risk of niche (RR 2.86, 95% CI 1.03 to 7.96; 2 studies, 97 women; I^2^ = 0.0%; low-certainty evidence; Figure S6). Further, Wang 2025 compared the risk of symptoms in niches with and without ectopic endometrium, finding a large increase in risk (RR 7.48, 95% CI 3.06 to 18.25; 1 study, 155 women; low-certainty evidence).

#### Asynchronous menstrual cycle

Three studies investigated asynchrony of the menstrual cycle, comparing cells in the niche to specimens of the uterine cavity. The estimated pooled prevalence was 30% (95% CI 13 to 55%; 3 studies, 157 women; I^2^ = 87.4%; very low-certainty evidence; Figure S7). Restricting the analysis to studies not at critical risk of bias yields a slightly higher estimate of 42% (95% CI 23 to 63%; 2 studies, 119 women; I^2^ = 80.9%; very low-certainty evidence).

#### Vascular remodeling

Vessels with prominent wall proliferation, referred to as thick-walled vessels, were investigated by six studies, observing a pooled prevalence of 81% (95% CI 75 to 86%; 6 studies, 205 women; I^2^ = 0.0%; low-certainty evidence; Figure S8).

#### Immunohistochemistry markers (inflammation and cell proliferation)

Four studies conducted immunohistochemistry analyses on markers of inflammation, inflammatory cells, cell proliferation, or hormonal expression (Tables S3 and S4). All studies, except for Wang 2025, compared these markers in women with niches with those without niches but with prior cesareans. Three studies assessed inflammatory markers in niche specimens, including CD138, CD16, CD3, CD20, CD56, CD68, CD163, myeloperoxidase (MPO), tryptase, and tumor necrosis factor-α (TNF-α). They reported increased expression of CD138, CD16, and TNF-α; decreased expression of CD3, CD20, CD68, CD163, and tryptase; and no significant change in CD56 and MPO (Table S3).

Regarding cell proliferation and hormonal receptors, three studies assessed markers such as α-smooth muscle actin (α-SMA), Ki67, CD10, CD31, estrogen receptor (ER), and progesterone receptor (PR). They reported significantly increased expression of CD31; α-SMA was either similar or increased; and Ki67, CD10, ER, and PR showed no significant change (Table S4).

### Certainty of evidence

As most of the publications consisted of retrospective observational studies, the certainty of our observations was low to very low. The domains in which we downgraded the quality of evidence were inconsistency (one level), as studies generally reported non-standardized findings, and risk of bias extracted from ROBINS-I (two levels). Therefore, the certainty started at high and was downgraded by three levels (very low-certainty) in some cases and by two levels when inconsistency was not substantial (low-certainty).

## DISCUSSION

### Summary of findings

In this systematic review of women diagnosed with uterine niches in the cesarean scar and presenting chiefly with infertility and abnormal uterine bleeding, the most prevalent histological findings were fibrosis in 83% (51% to 96%), endometrial defects in 88% (81% to 93%), thick-walled vessels in 81% (75% to 86%), and chronic inflammation in 54% (25% to 81%). Furthermore, compared to women with prior cesarean deliveries but without niches, there is higher endometritis (CD138), increased neutrophils (CD16), and TNF-α expression, especially around thick-walled vessels, while T lymphocytes (CD3), B lymphocytes (CD20), macrophages (CD68), M2 macrophages (CD163), and tryptase expression are comparatively reduced.

Although ectopic endometrium was not highly prevalent, women with uterine niches had nearly a threefold higher risk of ectopic endometrium compared to those with prior cesarean deliveries. A similar increase in risk was observed for endometrial defects. A less frequent but notable observation was menstrual cycle asynchrony between the niche and surrounding cells.

### Limitations of the findings

A key limitation was the unclear reporting of pathological findings. Certain histological features were described in some studies but not in others, raising uncertainty about whether these abnormalities were truly absent or simply not assessed. This ambiguity also raises concerns about the consistency of specimen evaluation, specifically, whether all samples were analyzed using the same criteria and by the same group of pathologists.

In addition, the high heterogeneity observed in our analyses represents an important limitation. While some variability was anticipated, given the unstandardized patient selection and inherent design limitations of the included studies, the extent of heterogeneity may also reflect true differences in patient factors (age and comorbidities), the inflammatory stage, and the degree of tissue remodeling. However, it remains unclear whether these variations stem from between-study bias or genuine biological differences in the inflammatory response across study populations.

One observation that supports the latter hypothesis is the apparent association between the prevalence of chronic inflammation and extensive fibrosis across studies. In some cases, fibrosis was highly prevalent while chronic inflammation was not reported, whereas in others, chronic inflammation was common and extensive fibrosis was less frequently observed.

### Immune-mediated inflammation

The extent of damage to the uterine wall, particularly the endometrium, may influence both the magnitude of the inflammatory response and the likelihood of its chronic persistence ^28,29^. Other factors, such as genetic predisposition, are also likely to play a role, although these remain poorly understood in uterine physiology.

Altogether, there is a plausible indication that immune-mediated inflammation is central to the pathogenesis of niches. Higuchi 2021 ^20^ observed a distinct phenotype of immune cell recruitment in patients with niches compared to those with previous cesarean deliveries. There is reduced expression of anti-inflammatory macrophages and increased markers of chronic inflammatory responses.

In comparison, a similar immune environment is observed in eutopic endometrial cells of women with adenomyosis, with upregulation of pro-inflammatory markers^30^. Persistence of inflammation is attributed to both altered hormonal expression and signaling from ectopic cells.

The initial disturbance leading to an altered immune microenvironment in uterine niches could be hypoxia caused by surgical injury or suture techniques. Increased expression of hypoxia-inducible factor 1α and consequential classic epithelial-mesenchymal changes has been reported in patients with endometriosis ^31^. Although reversal of these conditions can ultimately lead to the resolution of abnormal signaling, chronic expression of transforming growth factor (TGF)-β1 and long-term effects on tissue organization are possible ^32^. Resistance to cell death, in a dysfunctional state, for example, is evidenced by increased expression of TNF-α in these patients.

### Endometrial dysfunction

Structural defects in the eutopic endometrium were present in nearly all women, which might explain the ‘chronic wound’ behavior. After impaired cell-cell communication in the eutopic endometrium, there is an increased probability of disorganization and growth into the myometrium. Further, it has been proposed that the presence of ectopic endometrium and activation of TGF-β signaling increase local metabolic demand, trigger hypoxia signaling ^33^, and pro-angiogenic factors associated with inflammation ^34^. This could contribute to the development of remodeled vessels and increased α-SMA deposition observed in the defect area. Also, perivascular fibroblasts, activated by endothelial-derived signals, are prone to adopt a pro-inflammatory phenotype ^35^.

In healthy women, endometrial stromal cells’ responsiveness to estrogen promotes angiogenesis and increased capillary diameter and growth from the basalis layer. Injury to the endometrium causes an intracellular increase in COX-2 activity, and consequently of prostaglandins, of note, prostaglandin E2 (PGE2) ^36^. Thereafter, PGE2 increases cAMP activity, which mediates aromatase conversion of androgens. In persistent inflammation, estrogen interacts with estrogen alpha receptors on the adjacent myometrium, promoting muscle contraction, tissue auto-traumatization, and subverts conventional regenerative pathways ^37^.

Furthermore, altered vasculature, angiogenesis, and asynchrony could increase the propensity for bleeding within uterine niches. These areas of frequent bleeding lead to activation of the coagulation cascade and pro-inflammatory responses ^39^.

Secondary infertility is also common. A recent systematic review by Vitagliano and collaborators ^40^ demonstrated that infertility is more closely associated with the presence of uterine niches than with cesarean delivery itself. Such observation reinforces the view that adequately healed cesarean scars are generally not problematic and may only confer a very modest increase in risk. In contrast, poor scar healing, characterized by the formation of uterine niches, creates the necessary conditions for adverse reproductive outcomes.

Plausible mechanisms for niche-associated infertility include reduced endometrial receptivity during implantation due to dysfunctional signaling and embryotoxic effects of chronic inflammation ^6^. In particular, fibrosis may be associated with a defective or poorly responsive endometrium. This mirrors observations in other gynecological conditions, such as endometriosis, where extensive fibrosis is linked to impaired endometrial function and reduced implantation potential ^41^.

For both infertility and abnormal uterine bleeding, hysteroscopic resection results in restoration of normal functional endometrium and resolution or improvement of symptoms in approximately 70% of women ^25,26,42^. The underlying mechanism may involve re-epithelialization driven by healthy progenitor cells ^29^.

One study reported better bleeding control with levonorgestrel-releasing intrauterine devices in patients showing more clinical signs of vascular proliferation, suggesting that certain histological patterns might influence treatment efficacy ^43^. In theory, pronounced fibrosis results in decreased PGE2 activity, whereas substantial vascular remodeling could indicate higher underlying inflammation and hormonal imbalance.

## Data Availability

All data produced in the present study are available upon reasonable request to the authors

## FIGURE LEGENDS

**Figure S1 Legend** Prevalence of histological findings per study.

**Figure S2 Legend** Subgroup analysis based on the risk of bias of the proportion of chronic inflammation in histopathological observations in women with symptomatic uterine niches. IV, inverse variance.

**Figure S3 Legend** Subgroup analysis based on the risk of bias of the proportion of fibrosis in histopathological observations in women with symptomatic uterine niches. IV, inverse variance.

**Figure S4 Legend** Relative risk of endometrial defects in histopathological observations in women with symptomatic uterine niches compared to women with previous cesarean deliveries without uterine niches. IV, inverse variance.

**Figure S5 Legend** Proportion of ectopic endometrium in histopathological observations in women with symptomatic uterine niches. IV, inverse variance.

**Figure S6 Legend** Relative risk of ectopic endometrium in histopathological observations in women with symptomatic uterine niches compared to women with previous cesarean deliveries without uterine niches. IV, inverse variance.

**Figure S7 Legend** Proportion of asynchronous menstrual cycle in histopathological observations in women with symptomatic uterine niches. IV, inverse variance.

**Figure S8 Legend** Proportion of thick-walled vessels in histopathological observations in women with symptomatic uterine niches. IV, inverse variance.

